# Inferring Multi-Organ Genetic Causal Connections using Imaging and Clinical Data through Mendelian Randomization

**DOI:** 10.1101/2023.05.22.23290355

**Authors:** Juan Shu, Rong Zheng, Julio Chirinos, Carlos Copana, Bingxuan Li, Zirui Fan, Xiaochen Yang, Yilin Yang, Xiyao Wang, Yujue Li, Bowei Xi, Tengfei Li, Hongtu Zhu, Bingxin Zhao

## Abstract

Understanding the complex causal relationships among major clinical outcomes and the causal interplay among multiple organs remains a significant challenge. By using imaging phenotypes, we can characterize the functional and structural architecture of major human organs. Mendelian randomization (MR) provides a valuable framework for inferring causality by leveraging genetic variants as instrumental variables. In this study, we conducted a systematic multi-organ MR analysis involving 402 imaging traits and 372 clinical outcomes. Our analysis revealed 184 genetic causal links for 58 diseases and 56 imaging traits across various organs, tissues, and systems, including the brain, heart, liver, kidney, lung, pancreas, spleen, adipose tissue, and skeletal system. We identified intra-organ causal connections, such as the bidirectional genetic links between Alzheimer’s disease and brain function, as well as inter-organ causal effects, such as the impact of heart diseases on brain health. Metabolic disorders, such as diabetes, exhibited causal effects across multiple organs. These findings shed light on the genetic causal links spanning multiple organs, providing insights into the intricate relationships between organ functions and clinical outcomes.

---

Medical imaging techniques such as magnetic resonance imaging (MRI) provide important information about the structure and function of many human organs, such as the brain, heart, liver, and kidney. Their derived imaging traits have found widespread use in both clinical research and practical applications. For example, structural and functional imaging traits extracted from brain MRIs have consistently revealed abnormalities associated with Alzheimer’s disease, particularly within the hippocampal region^1^. Cardiovascular MRI (CMR) provides quantitative data regarding ventricular function, cardiovascular structure, and myocardial perfusion, all of which are intricately linked to cardiovascular diseases^2^. Furthermore, skeletal dual-energy X-ray absorptiometry (DXA) aids in identifying novel genetic variants that influence the human skeletal structure, thereby revealing significant evolutionary trends in human anatomical changes leading to pathogenesis^3^. Several large-scale organ imaging datasets (on the scale of over 10,000 participants) have recently been made publicly available, revealing details about human organ structure and function^4–8^. A large number of complex traits and clinical outcomes have been found to associate with organ imaging traits in these population-based studies^9,10^.

Despite these advances, inherent limitations in observational data pose challenges in definitively establishing causal relationships between imaging traits and clinical outcomes, as well as in gaining a comprehensive understanding of the causal interplay across various organs^11^. Mendelian randomization (MR) uses genetic variants as instrumental variables to infer causality from observational data^12,13^. Operating under certain assumptions regarding genetic, exposure, and outcome variables, MR is an epidemiological tool to uncover causal relationships between exposure and outcome variables, while controlling for confounding factors. Both family and population-based studies have demonstrated that numerous imaging traits and complex diseases are profoundly influenced by genetics. Hundreds of associated genetic loci have been pinpointed in large-scale genome-wide association studies (GWAS)^3,8,14–28^. Using these GWAS summary-level data (summary statistics), MR methods can unveil causal relationships between imaging measurements and clinical outcomes. Numerous recent MR studies have explored the genetic causality of imaging traits^11,29–34^. However, a common limitation of these MR studies is their focus on a single organ (or imaging modality) and/or a single disease, or diseases in a specific domain, such as brain imaging and psychiatric disorders^29^. It is crucial to note, however, that many diseases act as the causes and/or consequences of functional and structural changes in various organs of the human body^35,36^. Cross-organ analysis can elucidate the complexity of human physiology and holds great potential to improve our understanding of a multitude of diseases, ultimately enhancing our strategies for their diagnosis, treatment and prevention. Consequently, it is essential to conduct MR analysis from a multi-organ perspective to comprehend the clinical implications of imaging traits amidst the complex interrelationships of organ systems.

In this paper, we carried out a systematic two-sample MR analysis to explore the causal relationships between multi-organ imaging and clinical endpoints. We consolidated GWAS summary statistics from 402 multi-organ imaging traits (with an average sample size *n* ≈ 35,000) from the UK Biobank (UKB)^37^ study along with 372 clinical outcomes (304 with more than 10,000 cases and 68 more with at least 5000 cases) sourced from the FinnGen project^27^ (**Tables S1-S2**). Our focus was on three major brain MRI modalities: 1) 101 regional brain volumes^21^ from structural MRI (sMRI); 2) 110 diffusion tensor imaging (DTI) parameters^23^ from diffusion MRI (dMRI); and 3) 90 functional activity (amplitude^38^) and connectivity traits from functional MRI (fMRI)^25^. Furthermore, we incorporated 82 CMR traits extracted from short-axis, long-axis, and aortic cine cardiac MRI^39,40^. We also considered 11 abdominal MRI traits, measuring volume, fat, or iron content in seven organs and tissues^8^, and eight DXA imaging traits that gauged the lengths of all long bones and the width of the hip and shoulder^3^. We applied 8 MR methods^41–49^ to investigate bidirectional genetic causal links. The study design is presented in **Figure 1A**, while **Figure 1B** offers a high-level summary of our key findings. Additional details on these multi-organ imaging traits are provided in the **Methods** section.

**Fig. 1.**
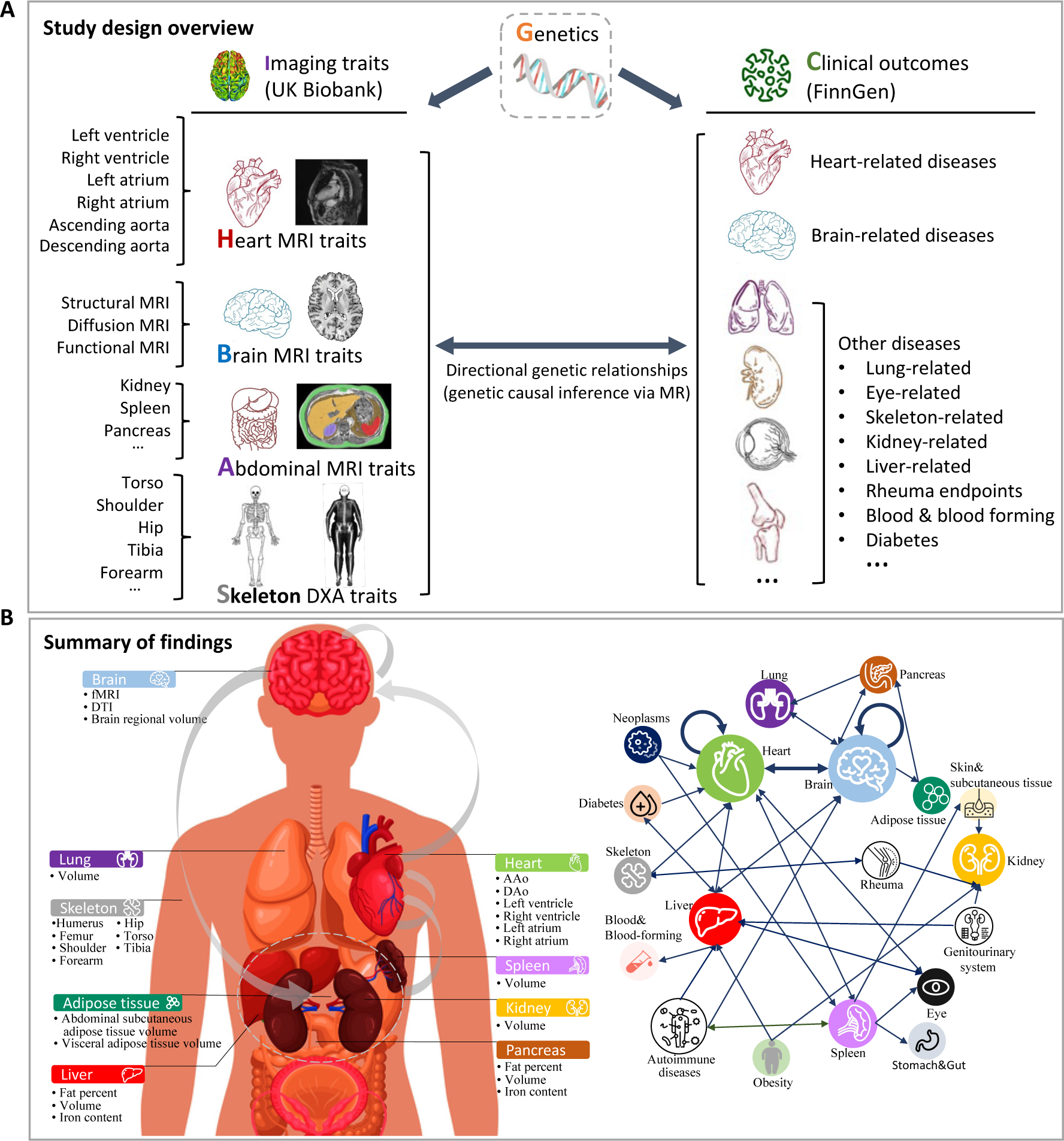
Overview of study design and findings. (**A**). An overview of our multi-organ imaging genetic study, investigating 372 clinical outcomes. We used diverse imaging traits including multi-modal brain, cardiac, abdominal, and skeletal DXA imaging to explore their relationships with the clinical endpoints. Our study encompasses a comprehensive range of brain imaging modalities, such as structural MRI, diffusion MRI, and resting-state fMRI. The cardiac imaging data comprise short-axis, long-axis, and aortic cine images. Volume, iron content, and percent fat were measured across 6 different abdominal organs and tissues, yielding in 11 image-derived abdominal phenotypes. Additionally, we included 8 skeleton imaging traits encompassing long bone lengths as well as hip and shoulder width. (**B**). A high-level summary of our bidirectional findings. The left panel displayed all the imaging traits that have been used in the study. The grey arrow demonstrates the main findings, such as the intra-brain, intra-heart, brain-heart, and brain-abdominal-organs causal connections. The right panel depicts the intricate web of causal interactions among various organs as uncovered in our study. The width of an arrow corresponds to the volume of findings associated with it.

## RESULTS

### Genetic causality between brain imaging and multi-organ diseases

We explored the causal relationship between brain imaging traits and multi-organ diseases. At a false discovery rate (FDR) 5% level (*P* < 1.31×10^−4^, adjusting for multiple testing for both directions), MR identified 33 significant genetic causal effects of 6 diseases (within 4 major categories) on 6 brain imaging traits. These categories included diseases of the nervous system, diseases of the circulatory system, diseases marked as autoimmune origin, and diseases of the digestive system. Among these 33 significant findings, 14 were associated with hypertension (9 with hypertension and 5 with essential hypertension), 12 with Alzheimer’s disease and dementia, 5 with autoimmune diseases, 1 with allergic asthma, and 1 with atrial fibrillation (**Table S3**).

Our results revealed that neurodegenerative conditions, including dementia and Alzheimer’s disease, could potentially impair functional activity across various networks. Specifically, dementia and Alzheimer’s disease were consistently found to compromise the integrity of the default mode (*β* < −0.09, *P* < 2.19×10^−5^, >6/8 MR methods significant), dorsal-attention (*β* < −0.05, *P* < 2.24×10^−5^, >5/8 MR methods significant), and frontoparietal networks (*β* < −0.03, *P* < 4.53×10^−5^, >5/8 MR methods significant) (**Fig. 2A**). In addition to degenerative neurological diseases, we consistently observed causal effects of hypertension on regional brain volume, fMRI traits, and DTI parameters. For example, hypertension had negative causal effects on grey matter volume (*β* < −0.03, *P* < 1.30×10^−10^, 5/8 MR methods significant). Similarly, a potential negative effect of hypertension was observed on the fractional anisotropy (FA) of the body of the corpus callosum tract (BCC, *β* < −0.07, *P* < 1.12×10^−6^, 6/8 MR methods significant) and the genu of the corpus callosum (GCC, *β* < −0.01, *P* < 2.29×10^−5^, 6/8 MR methods significant). Conversely, positive effects were found on other DTI parameters, such as axial diffusivity (AD), mean diffusivity (MD) and radial diffusivity (RD) of superior corona radiata (SCR), RD of BCC, as well as RD of GCC (*β* > 0.01, *P* < 4.30×10^−5^, 6/8 MR methods significant). It makes sense as lower MD and higher FA values typically indicating better white matter health^50^. In addition, we observed that hypertension was causally related to decreased functional connectivity of the orbito-affective network (*β* > −0.05, *P* < 1.16×10^−5^, 6/8 MR methods significant) (**Fig. 2B**). Autoimmune diseases were mostly found to affect DTI parameters, such as the AD and MD of SCR (*β* < −0.04, *P* < 9.43×10^−5^, >5/8 MR methods significant) (**Fig. 2C**).

**Fig. 2.**
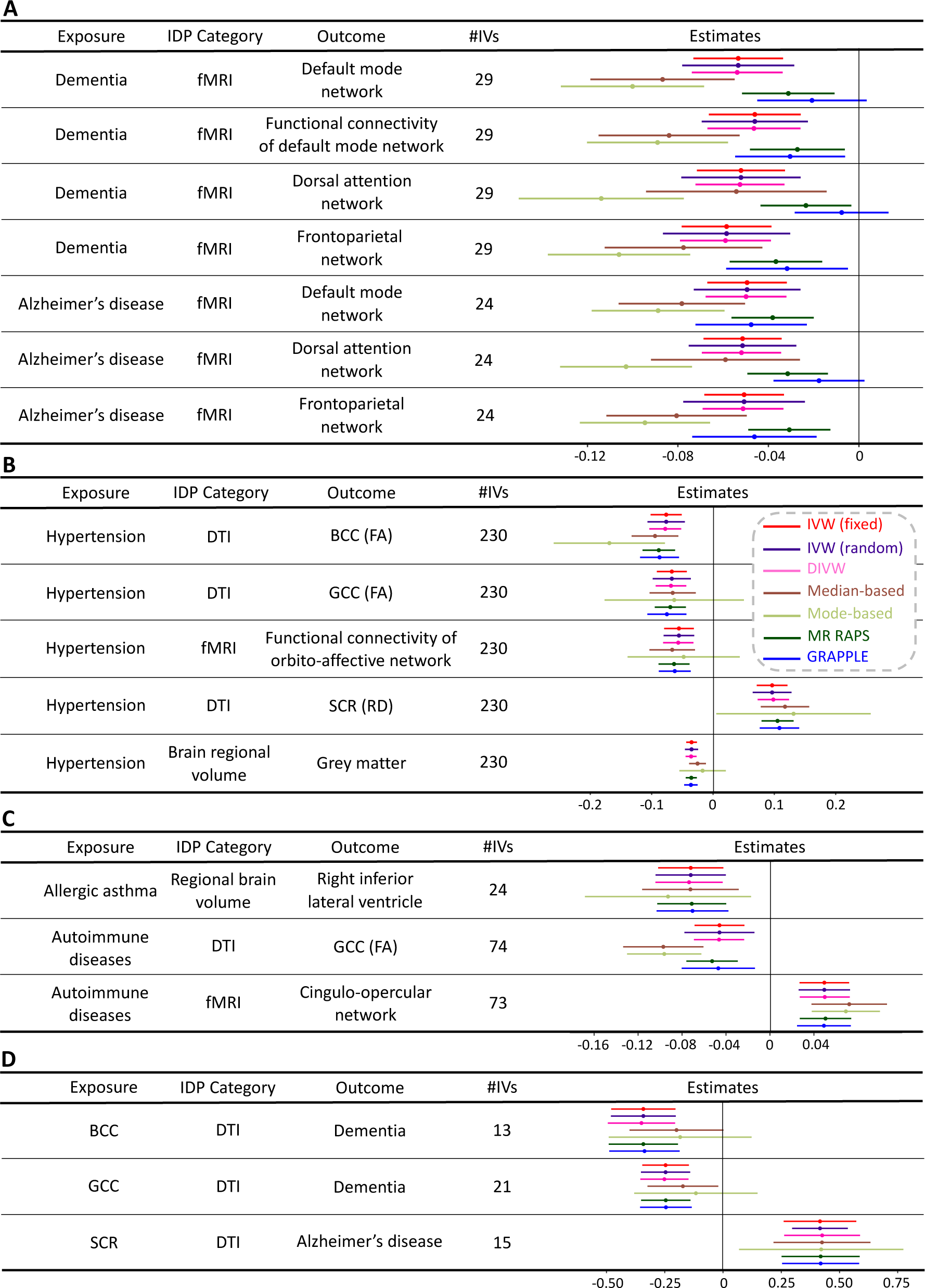
Selected genetic causal effects of clinical outcomes on brain imaging traits. We illustrated selected significant (*P* < 1.31×10^−4^) putative causal genetic links from clinical endpoints (Exposure) to brain imaging traits (Outcome), with adjustment for multiple testing using the FDR procedure. (**A**). The causal effect of degenerative neurological diseases on brain imaging traits. (**B**). The causal effect of hypertension on brain imaging traits. (**C**). The causal effect of other diseases on brain imaging traits. (**D**). The causal effect of brain imaging traits on degenerative neurological diseases. The term ‘IDP Category’ is used to signify the category of imaging traits, while ‘#IVs’ stands for the number of genetic variants utilized as instrumental variables. Various MR methods and their associated regression coefficients are marked with different colors for easy identification. See **Table S1** for data resources on clinical endpoints and **Table S2** for data resources on imaging traits.

Brain and other organ-related diseases may also be affected by structural or functional changes in the brain. To investigate this, we used brain imaging traits as exposure variables and clinical endpoints as the outcome variables. At an FDR 5% level (*P* < 1.31×10^−4^), we identified 12 significant relationships between 5 brain imaging traits and 3 types of clinical endpoints (**Table S3**). Most of the significant imaging-disease pairs were related to DTI parameters and the majority of the significant findings pertained to brain diseases (11/12). For example, we found that the RD of BCC and GCC was causally related to dementia (*β* < −0.31, *P* < 1.47×10^−5^, >5/8 MR methods significant), and the FA of SCR was associated with an increased risk of Alzheimer’s disease (*β* > 0.33, *P* < 6.24×10^−5^, 7/8 MR methods significant) (**Fig. 2D**).

### Causal genetic relationships between CMR traits and clinical outcomes

We assessed the potential causal effects from clinical endpoints to CMR measures of heart structure and function. We identified 137 significant results at an FDR 5% level (*P* < 6.95×10^−4^), including 44 unique CMR traits of the ascending aorta (AAo), descending aorta (Dao), left atrium (LA), right ventricle (RV), and left ventricle (LV). These significant causal effects were identified in 20 unique clinical endpoints of 5 categories: diseases of the circulatory system (10/20), endocrine, nutritional and metabolic diseases (6/20), mental and behavioral disorders (2/20), diseases of eye and adnexa (1/20), as well as neoplasms (ICD2 & ICD-O-3, 1/20). Among the 137 significant findings, 117 were associated with heart-related diseases (114 diseases of the circulatory system and 3 cardiac arrhythmias), 15 with endocrine, nutritional and metabolic diseases, and 3 with mental and behavioral disorders (**Table S4**).

Causal effects of hypertension were widely observed on many LV traits, such as global and regional^51^ myocardial-wall thickness (segments 1-16, *β* > 0.07, *P* < 6.44×10^−5^, 7/8 MR methods significant) and radial strain traits (segments 7-9 and 12-15, *β* > 0.06, *P* < 8.14×10^−5^, 7/8 MR methods significant). In addition to LV traits, hypertension also affected multiple other CMR traits. For example, it had negative causal impacts on Aao and Dao distensibility^52–54^ (*β* < −0.10, *P* < 2.11×10^−5^, 7/8 MR methods significant), as well as positive effects on AAo and Dao minimum/maximum areas (AAo_max_ and AAo_min_ areas, *β* > 0.10, *P* < 3.32×10^−6^, 7/8 MR methods significant). The causal effects were also observed in several LA traits, such as negative effects on the left atrium ejection fraction (*β* < −0.07, *P* < 4.45×10^−4^, 7/8 MR methods significant) as well as positive effects on left atrium minimum/maximum volumes (LA_min_ and LA_max_ volumes, *β* > 0.06, *P* < 4.72×10^−4^, 6/8 MR methods significant). Similar to hypertension, we found that cardiovascular diseases and hypertensive diseases had causal effects on various LV traits, such as regional myocardial-wall thickness, left ventricular myocardial mass, and regional radial strain. We further identified heart-related disorders that contributed to alterations in heart structure and function. Conditions such as atrial fibrillation and flutter, heart failure, and cardiac arrhythmias were primarily linked to LA traits, including negative effects on the left atrium ejection fraction (*β* < −0.07, *P* < 1.35×10^−5^, 7/8 MR methods significant) and positive effects on LA_min_ and LA_max_ volumes (*β* > 0.07, *P* < 1.08×10^−6^, 7/8 MR methods significant) (**Fig. 3A**).

**Fig. 3.**
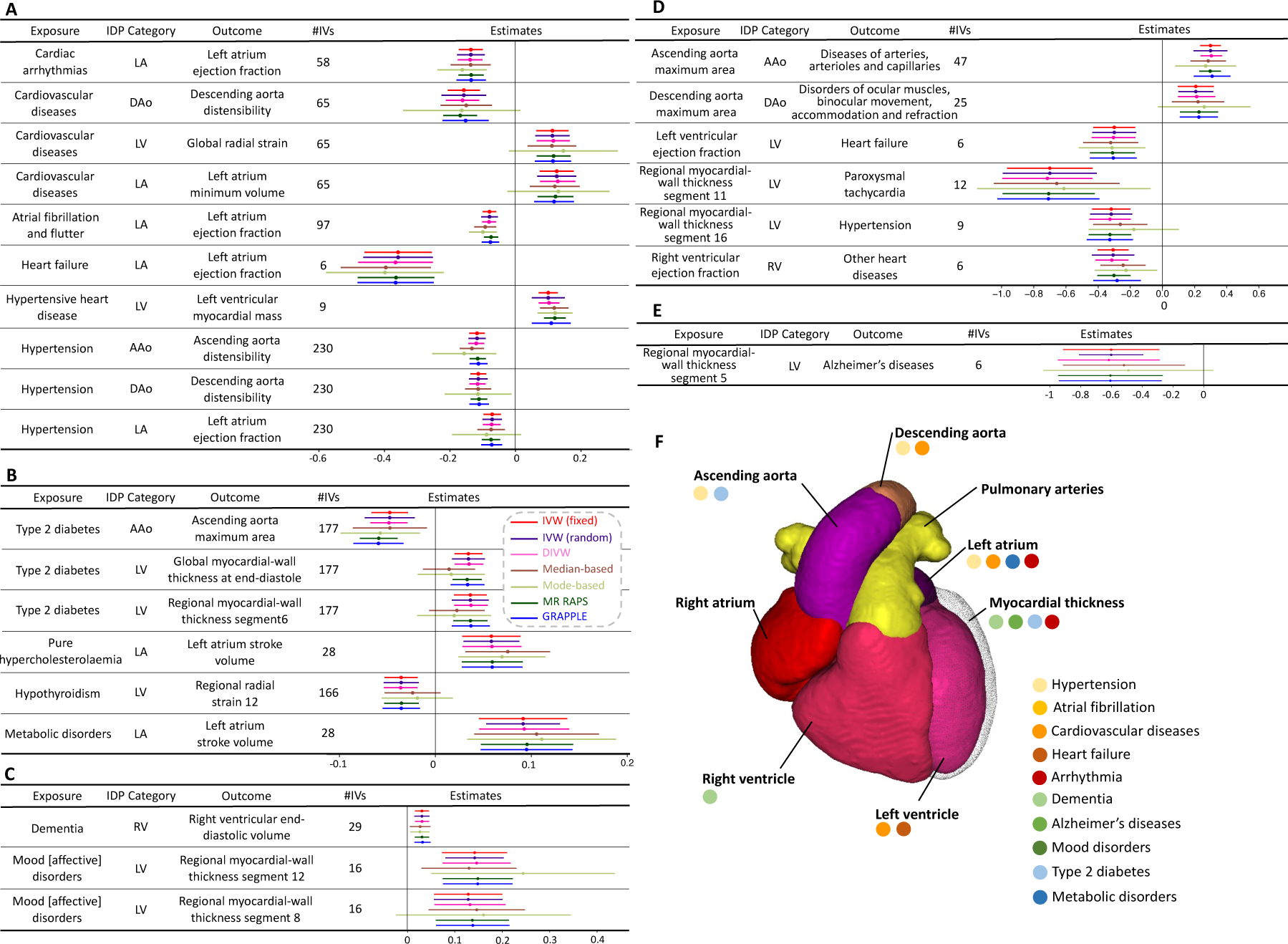
Selected genetic causal effects between heart imaging traits and clinical endpoints. (**A**). Selected significant (*P* < 6.95×10^−4^) causal genetic links from heart-related diseases (Exposure) to heart imaging traits (Outcome) with adjustment for multiple testing using the FDR procedure. (**B**). Selected significant (*P* < 6.95×10^−4^) causal genetic links from metabolic disorders (Exposure) to heart imaging traits (Outcome) with adjustment for multiple testing using the FDR procedure. (**C**). Selected significant (*P* < 6.95×10^−4^) causal genetic links from brain disorders (Exposure) to heart imaging traits (Outcome) with adjustment for multiple testing using the FDR procedure. (**D**). Selected significant (*P* < 6.95×10^−4^) causal genetic links from heart imaging traits (Exposure) to heart-related diseases (Outcome) with adjustment for multiple testing using the FDR procedure. (**E**). Selected significant (*P* < 6.95×10^−4^) causal genetic links from heart imaging traits (Exposure) to Alzheimer’s disease (Outcome) with adjustment for multiple testing using the FDR procedure. The term ‘IDP Category’ is used to signify the category of imaging traits, while ‘#IVs’ stands for the number of genetic variants utilized as instrumental variables. Various MR methods and their associated regression coefficients are marked with different colors for easy identification. (**F**). A high-level summary of our findings. See **Table S1** for data resources of clinical endpoints and **Table S2** for data resources on imaging traits.

We also observed causal effects of other diseases, such as endocrine, nutritional and metabolic disorders, on heart structure and function. For example, diabetes was observed to have negative effects on regional longitudinal strain (segment 2, |*β*|>0.03, *P* < 1.68×10^−4^, 5/8 MR methods significant) and positive effects on right ventricular ejection fraction (*β* > 0.04, *P* < 1.73×10^−4^, 5/8 MR methods significant). Additionally, type 2 diabetes led to a lower level of AAo_max_ and AAo_min_ areas (*β* < −0.04, *P* < 3.53×10^−4^, 5/8 MR methods significant) and larger global and regional myocardial-wall thickness (segments 5-6 and 10-12, *β* > 0.03, *P* < 5.35×10^−4^, 5/8 MR methods significant). Pure hypercholesterolemia and other metabolic disorders were causally linked to an enlarged left atrium stroke volume (*β* > 0.05, *P* < 3.31×10^−4^, 5/8 MR methods significant). Moreover, hypothyroidism negatively affected regional radial strain (segment 12, *β* < −0.05, *P* < 3.31×10^−4^, 5/8 MR methods significant) (**Fig. 3B**). It is also noteworthy that we found evidence suggestive of causality for brain disorders on heart phenotypes, such as a potential causal effect of dementia on right ventricular end-diastolic volume (*β* > 0.03, *P* < 2.42×10^−4^, 5/8 MR methods significant), as well as mood disorders on regional myocardial-wall thickness (segments 8 and 12, *β* < −0.12, *P* < 4.81×10^−4^, 5/8 MR methods significant) (**Fig. 3C**).

Structural and functional abnormalities within the heart may also increase the risk of multi-organ diseases, considering the role of the heart in pumping blood to all other organs^55^. We examined this direction by treating CMR traits as exposure variables and clinical endpoints as the outcomes. At an FDR 5% level (*P* < 6.95×10^−4^), we identified 20 significant causal pairs, with most related to heart-related diseases and a small portion to diseases of the nervous system. Some of these examples correspond to well-known relationships. For example, larger left ventricular ejection fraction was consistently found to reduce the risk of heart failure (*β* < −0.30, *P* < 2.38×10^−4^, 5/8 MR methods significant). (**Fig. 3D**). We also found that CMR traits, such as regional myocardial-wall thickness (segment 5), had causal effects on Alzheimer’s disease (*β* < −0.60, *P* < 4.85×10^−4^, 5/8 MR methods significant) (**Fig. 3E**).

### Causal genetic links between abdominal imaging traits and clinical outcomes

We first examined the causal effects of multi-organ diseases on abdominal imaging traits, including the volume or iron content of organs like the spleen, kidney, liver, lung, and pancreas^8^. At an FDR 5% level (*P* < 1.40×10^−3^), we identified 62 significant causal relationships between multi-organ diseases and abdominal imaging traits, after excluding straightforward associations such as the link between obesity and both adipose tissue volume and abdominal organ fat content. Notably, liver imaging traits constituted the majority of these findings (34/62). Brain disorders were the most prevalent among the significant relationships identified (23/62, mental and behavioral disorders (14) and diseases of the nervous system (9)), followed by endocrine, nutritional and metabolic diseases (18/62) (**Table S5**). These findings aligned with ongoing research on the complex interplay between the brain and abdominal organs, such as the brain-gut connection^56–58^, brain-kidney connection^59,60^, and brain-liver connection^61^. Specifically, Alzheimer’s disease and dementia were linked to a reduction in liver volume and percent of liver fat (|*β*|>0.02, *P* < 1.28×10^−4^, 6/8 MR methods significant). Sleep apnea was observed to positively influence the liver volume and the volume of abdominal subcutaneous adipose tissue (*β* > 0.19, *P* < 9.61×10^−4^, 6/8 MR methods significant). Furthermore, alcohol use disorder was associated with an increase in percent liver fat (*β* > 0.13, *P* < 2.09×10^−5^, 6/8 MR methods significant). In addition to the brain, we found that diabetes was causally related to a higher percent of liver fat (*β* > 0.04, *P* < 2.75×10^−5^, 5/8 MR methods significant) and increased pancreas fat (*β* > 0.5, *P* < 9.14×10^−5^, 5/8 MR methods significant). Furthermore, we found some autoimmune, rheumatic and infectious diseases were causally associated with lower levels of liver iron content (*β* < −0.04, *P* < 4.47×10^−4^, >6/8 MR methods significant) and larger spleen volume (*β* > 0.06, *P* < 4.47×10^−4^, 6/8 MR methods significant) (**Figs. 4A-C**).

**Fig. 4.**
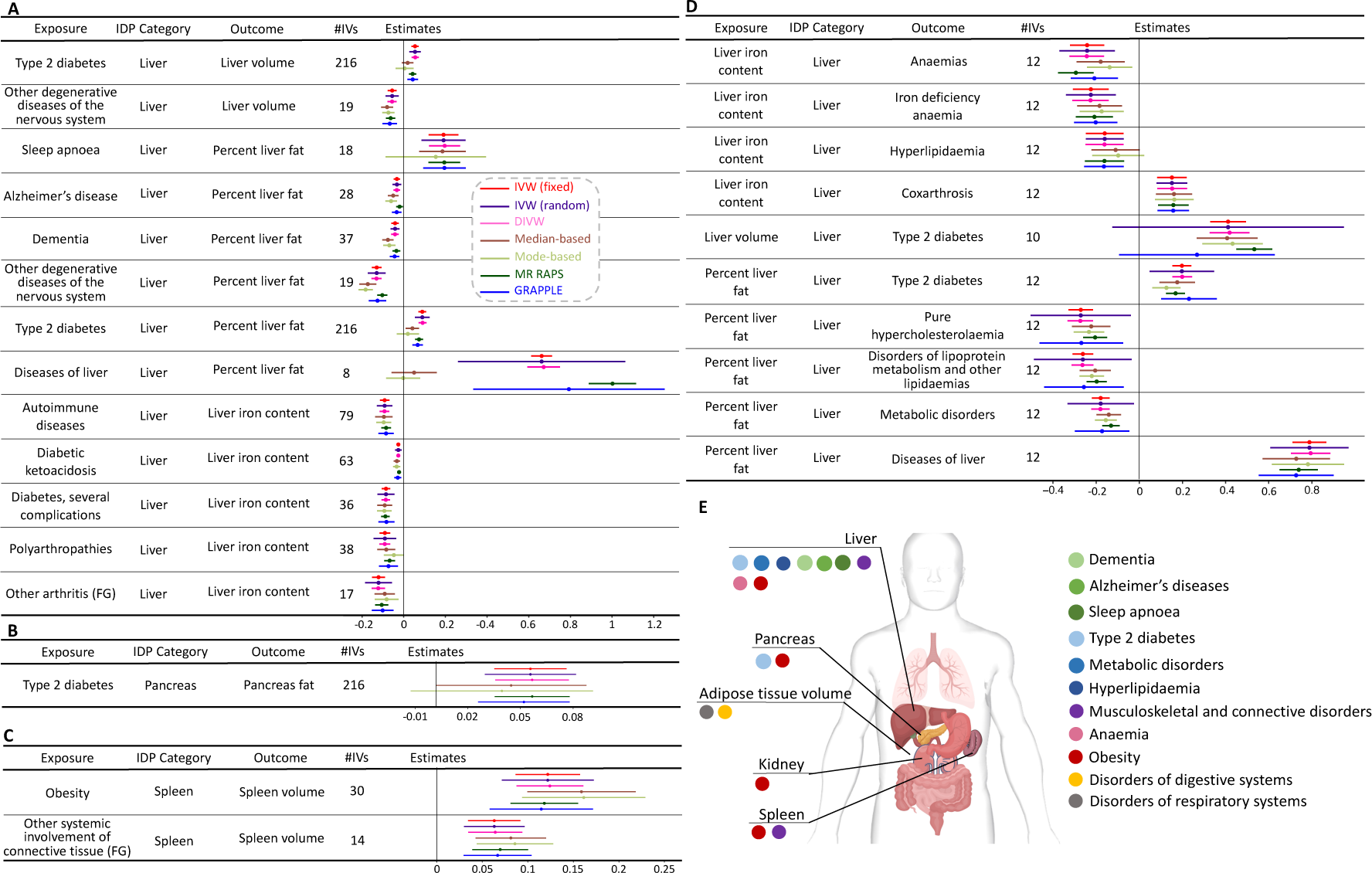
Selected genetic causal effects between abdominal imaging traits and clinical endpoints. **(A)** Selected significant (*P* < 1.40×10^−3^) causal genetic links from clinical endpoints (Exposure) to liver imaging traits (Outcome) with adjustment for multiple testing using the FDR procedure. (**B**). Selected significant (*P* < 1.40×10^−4^) causal genetic links from clinical endpoints (Exposure) to pancreas imaging traits (Outcome) with adjustment for multiple testing using the FDR procedure. (**C**). Selected significant (*P* < 1.40×10^−3^) causal genetic links from clinical endpoints (Exposure) to spleen imaging traits (Outcome) with adjustment for multiple testing using the FDR procedure. (**D**). Selected significant (*P* < 1.40×10^−4^) causal genetic links from liver imaging traits (Exposure) to pancreas clinical endpoints (Outcome) with adjustment for multiple testing using the FDR procedure. The term ‘IDP Category’ is used to signify the category of imaging traits, while ‘#IVs’ stands for the number of genetic variants utilized as instrumental variables. Various MR methods and their associated regression coefficients are marked with different colors for easy identification. (**E**). A high-level summary of our bidirectional findings. See **Table S1** for data resources on clinical endpoints and **Table S2** for data resources on imaging traits.

Next, we evaluated the opposite direction with abdominal imaging traits as exposure variables and multi-organ diseases as outcomes. At an FDR 5% level (*P* < 1.40×10^−3^), we identified 44 significant pairs, with heart-related diseases occupying 15 among them, followed by endocrine, nutritional metabolic disorders (10/44), disease of the respiratory system (4/44), blood and blood-forming (4/44), musculoskeletal and connective (4/44), and digestive system (4/44). The liver imaging traits were again the most frequent (35/44). For example, a higher percent of liver fat had causal effects on multiple heart-related diseases, such as angina pectoris, coronary angioplasty, coronary atherosclerosis, ischemic heart disease (wide definition), myocardial infarction (strict), as well as heart failure and coronary heart disease (*β* > 0.13, *P* < 1.40×10^−3^, 5/8 MR methods significant). Further, an enlarged spleen volume was linked to an increased risk of cardiovascular diseases, such as diseases of arteries, arterioles and capillaries (*β* > 0.25, *P* < 1.12×10^−3^, 5/8 MR methods significant). Beyond cardiovascular impacts, increased liver iron content significantly elevated the risk of anemias, particularly iron deficiency anemia (*β* > 0.19, *P* < 9.80×10^−4^, 5/8 MR methods significant). A causal relationship was observed between higher percent of liver fat and liver diseases (*β* > 0.13, *P* < 1.40×10^−3^, 8/8 MR methods significant), increased visceral adipose tissue volume and hernia of abdominal wall (*β* > 0.72, *P* < 1.40×10^−3^, 6/8 MR methods significant), as well as spleen volume and noninfective enteritis and colitis (*β* > 0.13, *P* < 9.66×10^−4^, 5/8 MR methods significant). Moreover, visceral adipose tissue volume led to an increased risk of asthma and chronic lower respiratory diseases (*β* > 0.30, *P* < 1.10×10^−3^, 5/8 MR methods significant) (**Fig. 4D**).

### Causal genetic links between skeleton DXA traits and clinical outcomes

The skeletal system serves as the foundational support for the human body, and as such, skeletal abnormalities could potentially contribute to the risk of multi-organ diseases. We identified 8 causal pairs at an FDR 5% level (*P* < 1.04×10^−4^), spanning conditions related to both the circulatory system and the musculoskeletal and connective tissue. Notably, an extended torso length was linked to an increased risk of several cardiovascular conditions, such as coronary atherosclerosis and myocardial infarction (|*β*|>18.72, *P* < 1.40×10^−3^, 5/8 MR methods significant). Furthermore, longer torso length was inked to several musculoskeletal and connective tissue disorders, such as internal derangement of the knee and meniscus derangement (|*β*|>21.20, *P* < 1.49×10^−5^, 5/8 MR methods significant) (**Fig. 5A**). We also uncovered potential causal effects of multiple organ diseases on DXA-derived skeleton traits. At an FDR 5% level (*P* < 1.04×10^−4^), we found 4 significant results that mainly on connective tissue disorders. For example, systemic connective tissue disorders and other systemic involvement of connective tissue were associated with lower average tibia height (|*β*|>0.0005, *P* < 3.61×10^−5^, 5/8 MR methods significant) (**Fig. 5B** and **Table S6**).

**Fig. 5.**
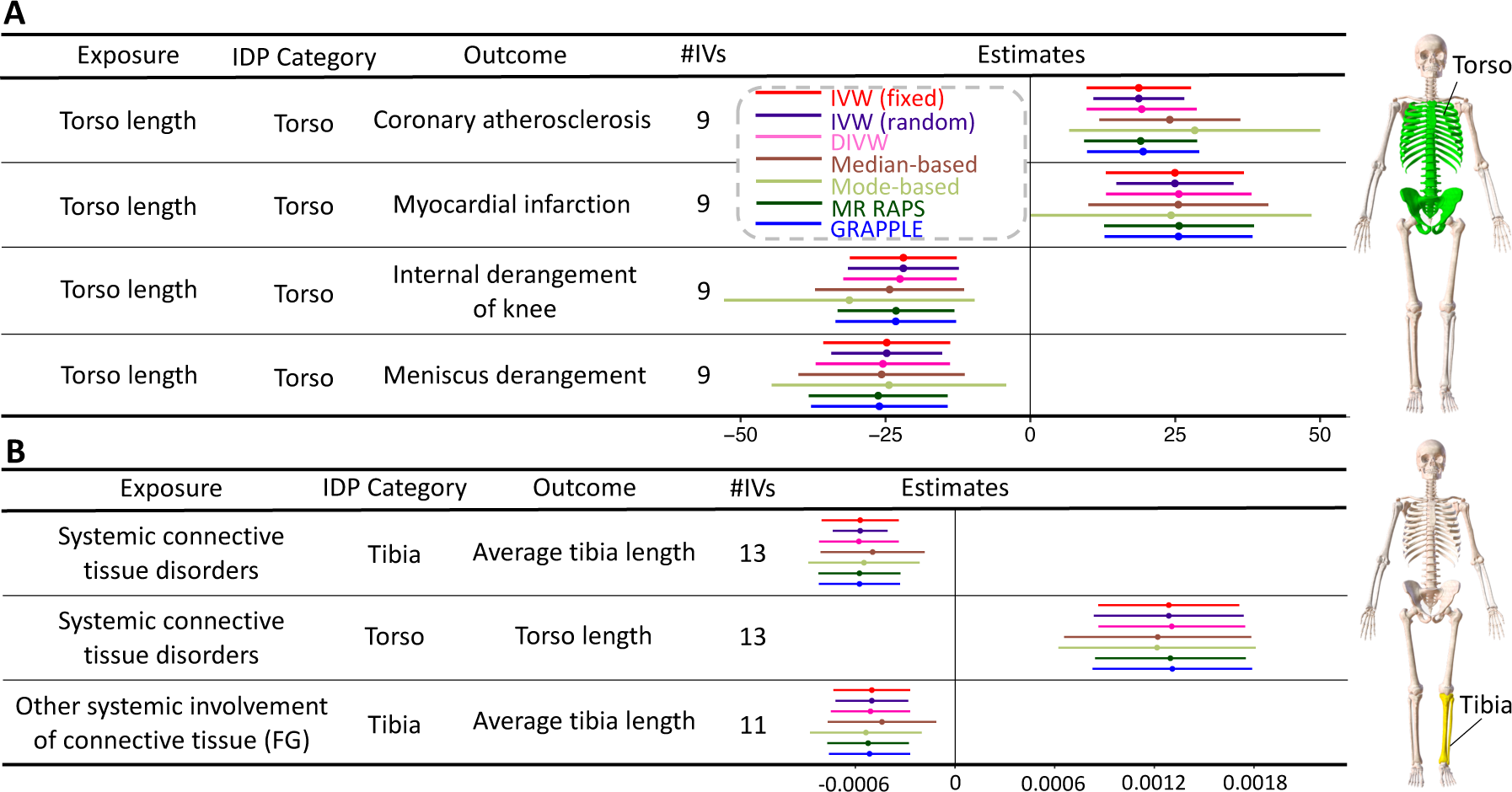
Selected genetic causal effects between skeleton imaging traits and clinical endpoints. We illustrated selected significant (*P* < 1.04×10^−4^) causal genetic links from (**A**) skeleton imaging traits (Exposure) to clinical endpoints (Outcome) with adjustment for multiple testing using the FDR procedure and (**B**) clinical endpoints (Exposure) to skeleton imaging traits (Outcome). The term ‘IDP Category’ is used to signify the category of imaging traits, while ‘#IVs’ stands for the number of genetic variants utilized as instrumental variables. Various MR methods and their associated regression coefficients are marked with different colors for easy identification. The skeleton diagram on the right shows the region of the corresponding IDP. See **Table S1** for data resources on clinical endpoints and **Table S2** for data resources on imaging traits.

## Discussion

Observational studies have identified numerous links between imaging-derived phenotypes and clinical outcomes. However, these associations are often influenced by residual confounding, complicating the accurate inference of causal effect sizes^62^. MR allows for the inference of causal relationships between exposure and outcome variables. MR leverages the natural and random assortment of genetic variants during meiosis, making these variants ideal instrumental variables for discerning causal effects. In the present study, we evaluated the causal links between 402 multi-organ imaging traits and clinical outcomes through bidirectional MR. To circumvent the issue of sample overlap^63^, which can bias the causal effect and has been overlooked in many current MR-based studies, we used a two-sample MR design and sourced our imaging and clinical data from different large-scale cohorts.

The interconnected nature of our organ systems suggested that diseases often affect more than one part of the human body^35,36,61,64^. The brain and heart are particularly critical, with the brain controlling a variety of functions, including reactions, emotions, vision, memory, and cognition^65,66^, while the heart acts pumps oxygen and nutrient-rich blood to other organs. Dysfunctions in various organs can potentially have detrimental effects on the brain and heart, and vice versa. Besides connections to the brain and heart, we also uncovered many other causal relationships across different organs. In this causal discovery research, we implemented stringent criteria for selecting significant causal pairs (**Methods**), with the goal being to ensure that all reported findings are statistically robust and capable of offering potential biological insights. Consequently, the absence of certain causal pairs in our study does not imply a lack of causal relationships. Moreover, our findings should be interpreted carefully in the context of known biology. Whereas some causal pairs are highly plausible and predictable based on current knowledge, many are novel and unexpected. These should stimulate further research to discern potential mechanistic pathways. A visualization of the interactions across different organ systems can be found in **Figure 1B**. We will discuss our findings in more detail below.

### Intra-brain causal connections

As expected, alterations in brain structure and function were found to be closely linked with brain disorders, with some of these relationships appearing to be bidirectional. We consistently identified causal connections between brain imaging traits and a variety of psychiatric disorders or neurological diseases, such as Alzheimer’s disease, dementia, mood disorder, and sleep apnea. For example, AD and dementia had an impact on fMRI traits, which were primarily observed as indicators of AD and dementia. Prior studies have consistently reported that resting fMRI connectivity patterns are disrupted in patients with Alzheimer’s disease^67,68^, particularly in brain regions involved in memory and cognitive function^69,70^. DTI abnormalities were found to be a precursor or an early sign of brain pathology that could lead to brain-related disorders^71,72^.

### Brain-heart causal connections

Despite the increasing number of association studies investigating the brain-heart interaction^40,73^, causal genetic links within these systems remain largely uncharted. In this study, we identified causal connections from hypertension to DTI parameters in multiple white matter tracts, including the SCR, BCC, and GCC. Additionally, hypertension had causal effects on regional brain volumes and fMRI traits, such as the mean activity within the orbito-affective network. These results could be attributed to the effects of hypertension on cerebral small vessel health^74,75^, leading to microvascular damage, which can disrupt white matter integrity, alter brain function observable in fMRI, and contribute to changes in brain structure, such as reduced regional volumes. Furthermore, elevated blood pressure may cause ischemia and impair the blood-brain barrier, contributing to neuroinflammation and neurodegeneration, reflected in these imaging traits^76–79^. Hence, effective management of hypertension through lifestyle adjustments and medication could help mitigate these adverse effects on the brain, which should be addressed in future studies.

We observed that heart structural measures, such as regional myocardial-wall thickness, exhibited potential causal effects on Alzheimer’s disease. Based on current knowledge, this connection does not seem to have clear underlying biologic plausibility, and should be interpreted cautiously until further knowledge is available regarding the role of cardiac abnormalities in the pathogenesis of Alzheimer’s disease, which is at present incompletely understood^80,81^. Causal connections were also detected from the brain to the heart. We found that mental or degenerative neurological disorders can affect CMR traits, such as mood disorder affecting regional myocardial-wall thickness. The reason behind it may be related to the stress response, which can lead to increased levels of stress hormones such as cortisol and adrenaline^82,83^. These hormones can cause an increase in blood pressure and heart rate, leading to hypertrophy (thickening) of the myocardial wall over time as the heart works harder to pump blood^84,85^. Mood disorders may also cause lifestyle changes (such as changes in dietary and physical activity patterns) that have deleterious effects on blood pressure and vascular function. Inflammation may also be involved, since mood disorders have been associated with increased levels of systemic inflammation, which is a known risk factor for various forms of cardiovascular disease^86,87^.

### Bidirectional connections between the brain and abdominal organs

Brain disorders were found to influence multiple abdominal organs. For example, we found that degenerative neurological diseases, such as Alzheimer’s disease and dementia may exert potential causal effects on several abdominal imaging traits, such as liver fat and volume, visceral/abdominal subcutaneous adipose tissue volume, as well as lung volume. Patients with Alzheimer’s disease and dementia often require multiple medications to manage symptoms, including antipsychotics, antidepressants, and medications aimed at slowing cognitive decline. Some of these medications can have hepatotoxic effects or interact with other drugs in ways that might impact liver function. Regular monitoring through liver function tests is important for patients on long-term medication regimens. Alzheimer’s disease and dementia also might lead to systemic inflammation^88^, which has been linked to abnormal intermediary metabolism, a key regulator of liver fat and function^89^. The connection between neurological disorders and adipose tissue volume might be related to reduced physical activity^90^ and corticosteroid medications^91^. The connection between Alzheimer’s disease and lung phenotypes may be related to effects of neurological diseases on respiratory muscle weakness^92,93^. This weakness could subsequently influence lung volume and functionality. Similar to Alzheimer’s disease and dementia, sleep apnea could also affect liver and adipose tissue. Whereas sleep apnea and liver/adipose tissue may have overlying common determinants, such as obesity, insulin resistance, decreased physical activity, and abnormal dietary patterns, sleep apnea may also impact various metabolic processes in its own right, which could contribute to further insulin resistance and fat accumulation in the liver. In addition to neurological diseases, our study revealed that alcohol use disorder can cause an increase in liver fat percentage. It is well documented that individuals suffering from alcohol use disorder are more prone to develop alcoholic fatty liver disease^94^.

Conversely, brain imaging traits were causally affected by various diseases of other organs or systems. For example, we found that asthma influenced regional brain volumes. Interestingly, prior studies have demonstrated that individuals with asthma may have diminished cognitive function, including impaired memory and attention, as well as changes in brain activity patterns during cognitive tasks^95,96^. Anxiety, stress, and depression, which are often experienced by individuals with asthma, can also result in alterations to brain structure. Notably, autoimmune diseases (as defined by FinnGen^27^) impacted brain imaging traits, such as DTI parameters of the SCR. Multiple sclerosis, an autoimmune disease affecting the central nervous system, can cause damage to the myelin sheath that surrounds axons and can occur in various brain regions^97,98^, including the internal capsule. This damage can disrupt neural connections passing through the anterior limb, leading to symptoms such as weakness, spasticity, and difficulty with balance and coordination. In rare autoimmune diseases, such as neuromyelitis optica^99^ and autoimmune encephalitis, inflammation, and damage can occur in the brain. The resulting neurological symptoms can vary based on the severity and location of the damage^100,101^. Additionally, autoimmune diseases causing systemic inflammation, such as rheumatoid arthritis and lupus, could potentially affect the brain^102^ and white matter tracts^103,104^. Chronic inflammation can lead to changes in the microstructure of white matter tracts, resulting in alterations in neural connectivity and function^105,106^.

### Intra-heart causal connections

Bidirectional causal relationships were observed between cardiovascular diseases and CMR traits. Hypertension was found to causally influence several CMR traits across different heart chambers and aorta regions. Conversely, alterations in CMR traits were contribute causally to heart diseases. These findings are well-aligned with our current understanding of the effects of hypertension on the heart. For example, hypertension can cause changes in the heart’s structure and function due to increased afterload, leading to left ventricular hypertrophy and changes in left atrial size,^107,108^ which can in turn contribute to the development of various complications, including atrial fibrillation, heart failure, and stroke^109^. Additionally, changes in cardiac structure might influence blood pressure regulation, since from a hemodynamic standpoint, blood pressure is the direct result of interactions between the cardiac pump and its coupled vascular load^110^.

### Causal connections between the heart and abdominal organs

Heart health was found to be causally linked with various multi-organ imaging traits or diseases. For example, liver fat was linked to multiple heart-related diseases, such as coronary atherosclerosis. This may be due to the key role of the liver in intermediary and lipoprotein metabolism, both of which are known contributors to the pathogenesis of atherosclerotic vascular disease. Additionally, heart failure was observed to cause an increase in spleen volume. The latter could be due to venous stasis and venous hypertension, which is a known consequence of heart failure. Indeed, increased venous pressure can lead to spleen enlargement, a condition known as splenomegaly^111^. Conversely, we found larger spleen volume to exhibit a causal effect on heart failure, a causal connection that is more difficult to interpret based on current knowledge, and which should stimulate further research. The pancreas also had causal effects on the cardiovascular system. For example, excess pancreas fat was associated with a higher risk of developing deep vein thrombosis of the lower extremities and pulmonary embolism. Pancreatic steatosis, a condition where fat accumulates in the pancreas, is linked with several metabolic abnormalities, including insulin resistance and inflammation, which can contribute to the development of cardiovascular diseases^112,113^.

### Causal connections between metabolic disorders and multiple organs

Diabetes was found to have an adverse impact on heart structural variations in AAo, RV, and LV. Multiple pathways may account for these causal associations, such as diabetic cardiomyopathy^114^, microvascular dysfunction^115,116^, and metabolic disturbances^117^. Diabetes can also affect the fat accumulation in the liver and pancreas. The liver plays a crucial role in managing glucose and fat metabolism. In the face of insulin resistance, the liver can accumulate fat, leading to non-alcoholic fatty liver disease. Insulin resistance prompts the liver to increase glucose production, contributing to high blood sugar levels, while also interfering with the liver’s ability to process fat, leading to fat accumulation. Fat accumulation is not limited to the liver; it can also occur in the pancreas. The pancreas is sensitive to the metabolic changes induced by insulin resistance and obesity. Excess fat within the pancreas may affect its function, particularly the insulin-producing beta cells. Studies suggest that fat accumulation in the pancreas can impair insulin secretion^118,119^, exacerbating the cycle of insulin resistance and beta-cell dysfunction seen in type 2 diabetes. Additionally, the excess fat (especially in the form of triglycerides) within the pancreas can lead to beta-cell dysfunction^120^. This dysfunction further impairs the body’s ability to produce and regulate insulin effectively, worsening glucose control^121–123^.

### Skeleton DXA traits

We found genetic causal connections between DXA-derived skeleton traits and multiple organ diseases. MR typically reflects long-term effects rather than immediate impacts, making it a highly suitable method to measure causal effects related to the skeletal system. This is because skeletal traits are less prone to rapid modifications over short periods. Skeletal traits were shown to be potentially causally affected by musculoskeletal and connective disorders, which might have multiple pathways, such as altered bone density^124^, abnormal bone growth and formation, as well as inflammatory conditions. Conversely, skeletal disorders could contribute to numerous organ diseases, with heart conditions being the most prevalent in our analysis. However, the precise underlying mechanisms of these potentially causal connection require further study.

### Limitations and conclusions

Our study has several limitations. First, our GWAS summary statistics were sourced from publicly available databases. Thus, we could not evaluate the impact of unobserved confounders, such as population stratification, on our results. Second, a common limitation that most existing MR methods share is that they require several model assumptions. This may result in model misspecifications and issues related to data heterogeneity when integrating data from different data resources^125^. We have systematically applied quality control measures and performed sensitivity analyses in our study. Future research implementing more advanced MR methods may relax some of these model assumptions^126,127^. Furthermore, MR studies are designed to examine the effects of lifetime exposure factors on outcomes, not interventions within a specified period. As a result, our findings may be interpreted differently than the rigorous results obtained from randomized controlled trials. Moreover, our findings have varying degrees of biologic plausibility based on current knowledge. They should therefore be interpreted accordingly, and reinterpreted as new mechanistic knowledge becomes available.

In conclusion, we used two-sample bidirectional MR analyses to comprehensively explore the multi-organ causal connections between 372 clinical outcomes and 402 image-derived phenotypes of various organ systems. Our results revealed robust genetic evidence supporting causal connections within and across multiple organs. These findings will be useful in understanding complex pathogenic mechanisms and will contribute to further mechanistic research and ultimately, to the early prediction and prevention of multi-organ diseases from a whole-body perspective.

## METHODS

Methods are available in the ***Methods*** section.

*Note: One supplementary table zip file are available*.

## Data Availability

We used summary-level GWAS data in this study, which can be obtained from the FinnGen project (https://www.finngen.fi/en/access_results), BIG-KP (https://bigkp.org/),
Heart-KP (https://heartkp.org/), and project-specific resources detailed in Liu., et al and Kun., et al. Our multi-organ MR results can be explored at https://mr4mo.org/

## ACKNOWLEDGEMENTS

Research reported in this publication was supported by the National Institute On Aging of the National Institutes of Health under Award Number RF1AG082938. The content is solely the responsibility of the authors and does not necessarily represent the official views of the National Institutes of Health. The study has also been partially supported by funding from the Wharton Dean’s Research Fund, Analytics at Wharton, and Purdue Statistics Department. Assistance for this project was also provided by the UNC Intellectual and Developmental Disabilities Research Center (NICHD; P50 HD103573). This research has been conducted using summary-level data from the UK Biobank study and the FinnGen research project. We would like to thank the individuals who represented themselves in the UK Biobank and FinnGen studies for their participation and the research teams for their efforts in collecting, processing, and disseminating these datasets. We would like to thank the research computing groups at the University of North Carolina at Chapel Hill, Purdue University, and the Wharton School of the University of Pennsylvania for providing computational resources and support that have contributed to these research results.

## AUTHOR CONTRIBUTIONS

J.S. and B.Z. designed the study. J.S., C.C., B.L., Z. F., X.Y., Y.Y, X.W., and Y.L. analyzed the data. R.Z. and J.C. helped interpret findings. B.X., T.L., and H.Z. provided feedback on the results. J.S. and B.Z. wrote the manuscript with feedback from all authors.

## METHODS

### Multi-organ imaging traits

The imaging data were sourced from the UK Biobank (UKB) study, which enrolled approximately 500,000 individuals aged between 40 and 69 from 2006 to 2010 (https://www.ukbiobank.ac.uk/). These multi-organ imaging data were collected from the ongoing UKB imaging study project (https://www.ukbiobank.ac.uk/explore-your-participation/contribute-further/imaging-study), which aims to collect brain, heart, and abdomen scans from 100,000 participants. Ethical approval for the UKB study was secured from the North West Multicentre Research Ethics Committee (approval number: 11/NW/0382).

Studies of brain and heart diseases usually rely on magnetic resonance imaging (MRI) scans, which are well-established clinical endophenotypes. Cardiovascular magnetic resonance imaging (CMR) is a set of MRI techniques that are designed to assess ventricular function, cardiovascular morphology, myocardial perfusion, and other cardiac functional and structural features^128,129^. They have been frequently used to reveal heart-related issues clinically. The CMR traits used in the paper were originally generated from the raw short-axis, long-axis, and aortic cine images using the state-of-the-art heart imaging segmentation and feature representation framework^39,130,131^. We divided the generated 82 CMR traits into 6 categories. The first two are aortic sections, namely ascending aorta (AAo) and descending aorta (DAo), which serve as the main ‘pipe’ in supplying blood to the entire body. The other four are the global measures of 4 cardiac chambers, including the left ventricle (LV), right ventricle (RV), left atrium (LA), and right atrium (RA), which altogether manage the heartbeat and blood flow. There are also some other traits, such as regional phenotypes of the left ventricle myocardial-wall thickness and strain (**Table S1**). The summary-level GWAS data of these 82 CMR traits were obtained from Zhao, et al. ^40^.

Brain MRI provides detailed information about brain structure and function^132^, such as abnormal growth, healthy aging, white matter diseases, structural issues, and functional abnormalities. In this paper, the summary-level GWAS data were collected from recent multi-modal image genetic studies, including regional brain volumes from structural MRI^21,133^ (sMRI), diffusion tensor imaging (DTI) parameters from diffusion MRI^23,134^ (dMRI), and functional activity (that is, amplitude^38^) and functional connectivity phenotypes from resting functional MRI^25^ (resting fMRI). In sMRI, we used ANTs^135^ to generate regional brain volumes for cortical and subcortical regions and global brain volume measures. In dMRI, we used the ENIGMA-DTI pipelines^136,137^ to generate tract-averaged parameters for fractional anisotropy, mean diffusivity, axial diffusivity, radial diffusivity, and mode of anisotropy in major white matter tracts and across the whole brain. For resting fMRI, we extracted phenotypes from brain parcellation-based analysis. We used the Glasser360 atlas^138^, which divided the cerebral cortex into 360 regions in 12 functional networks^139^. We considered 90 network-level resting fMRI phenotypes that evaluated interactions and spontaneous neural activity at rest.

The 11 imaging traits from abdominal MRI were derived by Liu., et al^8^ using deep learning methods in terms of volume, fat, and iron in several organs and tissues, such as the liver, spleen, kidney, lung, pancreas, and adipose tissue. Skeleton DXA traits, including all long bone lengths as well as hip and shoulder width, were derived by Kun, et al. ^3^ using deep learning methods on whole-body DXA images. All eight skeleton traits have been controlled for height.

### FinnGen clinical endpoints

We used 372 clinical endpoints (304 with more than 10,000 cases and 68 more with at least 5000 cases) collected by the FinnGen project, which were selected from the R9 release (https://www.finngen.fi/en/access_results). Our initial criterion for selecting diseases was to include those with more than 10,000 cases. For certain significant diseases, such as Alzheimer’s disease and neoplasm, we further relaxed the case number cutoff to 5,000. The 372 clinical endpoints covered diseases from various categories, namely, mental and behavioral disorders, diseases of the nervous system, diseases of the eye and adnexa, diseases of the genitourinary system, diseases of the circulatory systems, endocrine, nutritional and metabolic disorders, diseases marked as autoimmune origin, diseases of the respiratory system, diseases of the digestive system, diseases of blood and blood-forming, diseases of the musculoskeletal system and connective tissue, as well as some other endpoints. The definitions can be found at https://risteys.finregistry.fi/. The FinnGen data used in our study was obtained from separate cohorts than those supplying imaging traits, which were derived from the UKB study, thus ensuring there was no sample overlap. Detailed information of these 372 clinical variables can be found in **Table S2**.

### Mendelian randomization analysis

We examined the genetic causal relationships between the 402 imaging traits (101 brain regional volume traits, 110 brain DTI parameters, 90 network-level fMRI phenotypes, 82 CMR traits, 11 abdominal traits, and 8 skeleton DXA traits) and 372 clinical endpoints. Prior to conducting the Mendelian randomization (MR) analysis, we conducted standard preprocessing and quality control procedures. First, we selected genetic variants based on a significance threshold of 5×10^−8^ in the exposure GWAS data. To ensure the independence of the genetic variants used in MR, we implemented LD clumping with a window size of 10,000 and an r^2^ threshold of 0.01, using the 1000 Genomes European ancestry data as a reference panel. We used the TwoSampleMR package (https://mrcieu.github.io/TwoSampleMR/) for harmonization, which enabled us to accurately align alleles between the selected variants in the exposure and the reported effect on the outcome.

We assessed the performance of 8 MR methods, which included Inverse variance weighted (fixed effect), Inverse variance weighted (multiplicative random effect), MR-Egger, Simple Median, Weighted Median, Weighted Mode, DIVW, GRAPPLE, and MR-RAPS^41,42,44–49,140^, where MR Egger was used as the pleiotropy test and will not be included in the figures. To ensure the reliability of our results, we implemented several quality control procedures. We excluded causal estimates that relied on fewer than 6 genetic variants, as a larger number of genetic variants increases the statistical power of MR analysis^47,48^. We retained causal pairs that were significant in at least two out of the eight methods. We also screened for pleiotropy by using the MR-Egger intercept, the most used method for testing the pleiotropy assumption. If a causal estimate failed the MR-Egger intercept test, we required that it have significant results in at least one of the robust MR methods, such as Weighted Median, Weighted Model, MR-RAPS, or GRAPPLE. Additionally, we require all the selected causal pairs to be significant in more than half of the methods.

### Code availability

We made use of publicly available software and tools. Our analysis code will be made freely available at Zenodo.

### Data availability

We used summary-level GWAS data in this study, which can be obtained from the FinnGen project (https://www.finngen.fi/en/access_results), BIG-KP (https://bigkp.org/), Heart-KP (https://heartkp.org/), and project-specific resources are detailed in Liu., et al ^8^ and Kun, et al. ^3^.

## REFERENCES

1. Buckner, R.L., et al. Molecular, structural, and functional characterization of Alzheimer’s disease: evidence for a relationship between default activity, amyloid, and memory. Journal of neuroscience 25, 7709–7717 (2005).

2. Pennell, D.J., et al. Clinical indications for cardiovascular magnetic resonance (CMR): Consensus Panel report. European heart journal 25, 1940–1965 (2004).

3. Kun, E., et al. The genetic architecture and evolution of the human skeletal form. Science 381, eadf8009 (2023).

4. Petersen, S.E., et al. UK Biobank’s cardiovascular magnetic resonance protocol. Journal of cardiovascular magnetic resonance 18, 1–7 (2015).

5. Littlejohns, T.J., Sudlow, C., Allen, N.E. & Collins, R. UK Biobank: opportunities for cardiovascular research. European heart journal 40, 1158–1166 (2019).

6. Miller, K.L., et al. Multimodal population brain imaging in the UK Biobank prospective epidemiological study. Nature Neuroscience 19, 1523–1536 (2016).

7. Thompson, P.M., et al. ENIGMA and global neuroscience: A decade of large-scale studies of the brain in health and disease across more than 40 countries. Translational psychiatry 10, 1–28 (2020).

8. Liu, Y., et al. Genetic architecture of 11 organ traits derived from abdominal MRI using deep learning. Elife 10, e65554 (2021).

9. Smith, S.M. & Nichols, T.E. Statistical challenges in “big data” human neuroimaging. Neuron 97, 263–268 (2018).

10. Tian, Y.E., et al. Heterogeneous aging across multiple organ systems and prediction of chronic disease and mortality. Nature Medicine 29, 1221–1231 (2023).

11. Taschler, B., Smith, S.M. & Nichols, T.E. Causal inference on neuroimaging data with Mendelian randomisation. NeuroImage, 119385 (2022).

12. Sanderson, E., et al. Mendelian randomization. Nature Reviews Methods Primers 2, 1–21 (2022).

13. Pingault, J.-B., et al. Using genetic data to strengthen causal inference in observational research. Nature Reviews Genetics 19, 566–580 (2018).

14. Aung, N., et al. Genome-wide analysis of left ventricular image-derived phenotypes identifies fourteen loci associated with cardiac morphogenesis and heart failure development. Circulation 140, 1318–1330 (2019).

15. Córdova-Palomera, A., et al. Cardiac Imaging of Aortic Valve Area From 34 287 UK Biobank Participants Reveals Novel Genetic Associations and Shared Genetic Comorbidity With Multiple Disease Phenotypes. Circulation: Genomic and Precision Medicine 13, e003014 (2020).

16. Meyer, H.V., et al. Genetic and functional insights into the fractal structure of the heart. Nature 584, 589–594 (2020).

17. Pirruccello, J.P., et al. Analysis of cardiac magnetic resonance imaging in 36,000 individuals yields genetic insights into dilated cardiomyopathy. Nature communications 11, 1–10 (2020).

18. Pirruccello, J.P., et al. Genetic analysis of right heart structure and function in 40,000 people. Nature genetics 54, 792–803 (2022).

19. Thanaj, M., et al. Genetic and environmental determinants of diastolic heart function. Nature cardiovascular research 1, 361–371 (2022).

20. Elliott, L.T., et al. Genome-wide association studies of brain imaging phenotypes in UK Biobank. Nature 562, 210–216 (2018).

21. Zhao, B., et al. Genome-wide association analysis of 19,629 individuals identifies variants influencing regional brain volumes and refines their genetic co-architecture with cognitive and mental health traits. Nature genetics 51, 1637–1644 (2019).

22. Smith, S.M., et al. An expanded set of genome-wide association studies of brain imaging phenotypes in UK Biobank. Nature neuroscience 24, 737–745 (2021).

23. Zhao, B., et al. Common genetic variation influencing human white matter microstructure. Science 372, eabf3736 (2021).

24. Grasby, K.L., et al. The genetic architecture of the human cerebral cortex. Science 367, eaay6690 (2020).

25. Zhao, B., et al. Genetic influences on the intrinsic and extrinsic functional organizations of the cerebral cortex. medRxiv, 2021.2007. 2027.21261187 (2021).

26. Watanabe, K., et al. A global overview of pleiotropy and genetic architecture in complex traits. Nature genetics 51, 1339–1348 (2019).

27. Kurki, M.I., et al. FinnGen provides genetic insights from a well-phenotyped isolated population. Nature 613, 508–518 (2023).

28. Flynn, B.I., et al. Deep learning based phenotyping of medical images improves power for gene discovery of complex disease. medRxiv, 2023.2003. 2007.23286909 (2023).

29. Guo, J., et al. Mendelian randomization analyses support causal relationships between brain imaging-derived phenotypes and risk of psychiatric disorders. Nature Neuroscience 25, 1519–1527 (2022).

30. Chen, X., et al. Kidney damage causally affects the brain cortical structure: a Mendelian randomization study. EBioMedicine 72, 103592 (2021).

31. Williams, J.A., et al. Inflammation and brain structure in schizophrenia and other neuropsychiatric disorders: a Mendelian randomization study. JAMA psychiatry 79, 498–507 (2022).

32. Topiwala, A., et al. Associations between moderate alcohol consumption, brain iron, and cognition in UK Biobank participants: observational and mendelian randomization analyses. PLoS medicine 19, e1004039 (2022).

33. Holmes, M.V., et al. Mendelian randomization of blood lipids for coronary heart disease. European heart journal 36, 539–550 (2015).

34. Lamina, C. & Kronenberg, F. Estimation of the required lipoprotein (a)-lowering therapeutic effect size for reduction in coronary heart disease outcomes: a Mendelian randomization analysis. JAMA cardiology 4, 575–579 (2019).

35. Skou, S.T., et al. Multimorbidity. Nature Reviews Disease Primers 8, 48 (2022).

36. Langenberg, C., Hingorani, A.D. & Whitty, C.J. Biological and functional multimorbidity—from mechanisms to management. Nature Medicine 29, 1649–1657 (2023).

37. Sudlow, C., et al. UK biobank: an open access resource for identifying the causes of a wide range of complex diseases of middle and old age. PLoS medicine 12, e1001779 (2015).

38. Bijsterbosch, J., et al. Investigations into within-and between-subject resting-state amplitude variations. Neuroimage 159, 57–69 (2017).

39. Bai, W., et al. A population-based phenome-wide association study of cardiac and aortic structure and function. Nature Medicine 26, 1654–1662 (2020).

40. Zhao, B., et al. Heart-brain connections: Phenotypic and genetic insights from magnetic resonance images. Science 380, abn6598 (2023).

41. Bowden, J., et al. A framework for the investigation of pleiotropy in two-sample summary data Mendelian randomization. Statistics in medicine 36, 1783–1802 (2017).

42. Burgess, S., Butterworth, A. & Thompson, S.G. Mendelian randomization analysis with multiple genetic variants using summarized data. Genetic epidemiology 37, 658–665 (2013).

43. Bowden, J., et al. Improving the accuracy of two-sample summary-data Mendelian randomization: moving beyond the NOME assumption. Int J Epidemiol 48, 728–742 (2019).

44. Bowden, J., Davey Smith, G. & Burgess, S. Mendelian randomization with invalid instruments: effect estimation and bias detection through Egger regression. International journal of epidemiology 44, 512–525 (2015).

45. Hartwig, F.P., Davey Smith, G. & Bowden, J. Robust inference in summary data Mendelian randomization via the zero modal pleiotropy assumption. International journal of epidemiology 46, 1985–1998 (2017).

46. Bowden, J., Davey Smith, G., Haycock, P.C. & Burgess, S. Consistent estimation in Mendelian randomization with some invalid instruments using a weighted median estimator. Genetic epidemiology 40, 304–314 (2016).

47. Ye, T., Shao, J. & Kang, H. Debiased inverse-variance weighted estimator in two-sample summary-data Mendelian randomization. The Annals of statistics 49, 2079–2100 (2021).

48. Zhao, Q., Wang, J., Hemani, G., Bowden, J. & Small, D.S. Statistical inference in two-sample summary-data Mendelian randomization using robust adjusted profile score. The Annals of Statistics 48, 1742–1769 (2020).

49. Wang, J., et al. Causal inference for heritable phenotypic risk factors using heterogeneous genetic instruments. PLoS genetics 17, e1009575 (2021).

50. Bennett, I.J., Madden, D.J., Vaidya, C.J., Howard, D.V. & Howard Jr, J.H. Age-related differences in multiple measures of white matter integrity: A diffusion tensor imaging study of healthy aging. Human brain mapping 31, 378–390 (2010).

51. Segmentation, A.H.A.W.G.o.M., et al. Standardized myocardial segmentation and nomenclature for tomographic imaging of the heart: a statement for healthcare professionals from the Cardiac Imaging Committee of the Council on Clinical Cardiology of the American Heart Association. Circulation 105, 539–542 (2002).

52. Stratos, C., Stefanadis, C., Kallikazaros, I., Boudoulas, H. & Toutouzas, P. Ascending aorta distensibility abnormalities in hypertensive patients and response to nifedipine administration. The American journal of medicine 93, 505–512 (1992).

53. Asmar, R., et al. Aortic distensibility in normotensive, untreated and treated hypertensive patients. Blood pressure 4, 48–54 (1995).

54. Nabati, M., Namazi, S.S., Yazdani, J. & Sharif Nia, H. Relation between aortic stiffness index and distensibility with age in hypertensive patients. International journal of general medicine, 297–303 (2020).

55. Berman, M.N., Tupper, C. & Bhardwaj, A. Physiology, Left Ventricular Function. (2019).

56. Kim, D.-Y. & Camilleri, M. Serotonin: a mediator of the brain–gut connection. Official journal of the American College of Gastroenterology*| ACG* 95, 2698–2709 (2000).

57. Jones, M., Dilley, J., Drossman, D. & Crowell, M. Brain–gut connections in functional GI disorders: anatomic and physiologic relationships. Neurogastroenterology & Motility 18, 91–103 (2006).

58. Keefer, L., et al. A Rome working team report on brain-gut behavior therapies for disorders of gut-brain interaction. Gastroenterology 162, 300–315 (2022).

59. Xie, Z., Tong, S., Chu, X., Feng, T. & Geng, M. Chronic kidney disease and cognitive impairment: The kidney-brain axis. Kidney Diseases 8, 275–285 (2022).

60. de Donato, A., Buonincontri, V., Borriello, G., Martinelli, G. & Mone, P. The dopamine system: insights between kidney and brain. Kidney and Blood Pressure Research 47, 493–505 (2022).

61. McCracken, C., et al. Multi-organ imaging demonstrates the heart-brain-liver axis in UK Biobank participants. Nature Communications 13, 7839 (2022).

62. Walker, V.M., Zheng, J., Gaunt, T.R. & Smith, G.D. Phenotypic Causal Inference Using Genome-Wide Association Study Data: Mendelian Randomization and Beyond. Annu Rev Biomed Data Sci 5, 1–17 (2022).

63. Burgess, S., Davies, N.M. & Thompson, S.G. Bias due to participant overlap in two-sample Mendelian randomization. Genetic epidemiology 40, 597–608 (2016).

64. Jaggi, A., et al. A structural heart-brain axis mediates the association between cardiovascular risk and cognitive function. Imaging Neuroscience 2, 1–18 (2024).

65. Berridge, K.C. & Kringelbach, M.L. Pleasure systems in the brain. Neuron 86, 646–664 (2015).

66. Bressler, S.L. & Menon, V. Large-scale brain networks in cognition: emerging methods and principles. Trends Cogn Sci 14, 277–290 (2010).

67. Wang, K., et al. Altered functional connectivity in early Alzheimer’s disease: A resting-state fMRI study. Human brain mapping 28, 967–978 (2007).

68. Sorg, C., et al. Selective changes of resting-state networks in individuals at risk for Alzheimer’s disease. Proceedings of the National Academy of Sciences 104, 18760–18765 (2007).

69. Ranasinghe, K.G., et al. Regional functional connectivity predicts distinct cognitive impairments in Alzheimer’s disease spectrum. NeuroImage: Clinical 5, 385–395 (2014).

70. Pini, L., et al. A low-dimensional cognitive-network space in Alzheimer’s disease and frontotemporal dementia. Alzheimer’s Research & Therapy 14, 199 (2022).

71. Torso, M., et al. In vivo cortical diffusion imaging relates to Alzheimer’s disease neuropathology. Alzheimers Res Ther 15, 165 (2023).

72. Tu, M.-C., et al. Joint diffusional kurtosis magnetic resonance imaging analysis of white matter and the thalamus to identify subcortical ischemic vascular disease. Scientific Reports 14, 2570 (2024).

73. Liu, W., et al. Brain–heart communication in health and diseases. Brain Research Bulletin (2022).

74. Walker, K.A., Power, M.C. & Gottesman, R.F. Defining the relationship between hypertension, cognitive decline, and dementia: a review. Current hypertension reports 19, 1–16 (2017).

75. Zhang, H., et al. Reduced regional gray matter volume in patients with chronic obstructive pulmonary disease: a voxel-based morphometry study. American Journal of Neuroradiology 34, 334–339 (2013).

76. Yang, C., Hawkins, K.E., Doré, S. & Candelario-Jalil, E. Neuroinflammatory mechanisms of blood-brain barrier damage in ischemic stroke. American Journal of Physiology-Cell Physiology 316, C135–C153 (2019).

77. Carnevale, D., et al. Role of neuroinflammation in hypertension-induced brain amyloid pathology. Neurobiology of aging 33, 205. e219–205. e229 (2012).

78. Haspula, D. & Clark, M.A. Neuroinflammation and sympathetic overactivity: Mechanisms and implications in hypertension. Autonomic Neuroscience 210, 10–17 (2018).

79. Sweeney, M.D., Sagare, A.P. & Zlokovic, B.V. Blood–brain barrier breakdown in Alzheimer disease and other neurodegenerative disorders. Nature Reviews Neurology 14, 133–150 (2018).

80. Niedermeyer, E. Alzheimer disease: caused by primary deficiency of the cerebral blood flow. Clinical EEG and neuroscience 37, 175–177 (2006).

81. Kisler, K., Nelson, A.R., Montagne, A. & Zlokovic, B.V. Cerebral blood flow regulation and neurovascular dysfunction in Alzheimer disease. Nature Reviews Neuroscience 18, 419–434 (2017).

82. Chu, B., Marwaha, K., Sanvictores, T. & Ayers, D. Physiology, stress reaction. In StatPearls [Internet] (StatPearls Publishing, 2021).

83. Charmandari, E., Tsigos, C. & Chrousos, G. Endocrinology of the stress response. Annu. Rev. Physiol. 67, 259–284 (2005).

84. Colao, A., Marzullo, P., Di Somma, C. & Lombardi, G. Growth hormone and the heart. Clinical endocrinology 54, 137–154 (2001).

85. Fazio, S., et al. Growth hormone and heart performance: a novel mechanism of cardiac wall stress regulation in humans. European Heart Journal 18, 340–347 (1997).

86. Black, P.H. & Garbutt, L.D. Stress, inflammation and cardiovascular disease. Journal of psychosomatic research 52, 1–23 (2002).

87. Libby, P. Inflammation and cardiovascular disease mechanisms. The American journal of clinical nutrition 83, 456S–460S (2006).

88. Holmes, C. Systemic inflammation and A lzheimer’s disease. Neuropathology and applied neurobiology 39, 51–68 (2013).

89. Laleman, W., Claria, J., Van der Merwe, S., Moreau, R. & Trebicka, J. Systemic inflammation and acute-on-chronic liver failure: too much, not enough. Canadian Journal of Gastroenterology and Hepatology 2018(2018).

90. Scherder, E.J., Bogen, T., Eggermont, L.H., Hamers, J.P. & Swaab, D.F. The more physical inactivity, the more agitation in dementia. Int Psychogeriatr 22, 1203–1208 (2010).

91. Peckett, A.J., Wright, D.C. & Riddell, M.C. The effects of glucocorticoids on adipose tissue lipid metabolism. Metabolism 60, 1500–1510 (2011).

92. Polkey, M.I., Lyall, R.A., Moxham, J. & Leigh, P.N. Respiratory aspects of neurological disease. *Journal of Neurology*, Neurosurgery & Psychiatry 66, 5–15 (1999).

93. Pollock, R.D., Rafferty, G.F., Moxham, J. & Kalra, L. Respiratory muscle strength and training in stroke and neurology: a systematic review. International Journal of Stroke 8, 124–130 (2013).

94. Kushner, T. & Cafardi, J. Chronic liver disease and COVID-19: alcohol use disorder/alcohol-associated liver disease, nonalcoholic fatty liver disease/nonalcoholic steatohepatitis, autoimmune liver disease, and compensated cirrhosis. Clinical Liver Disease 15, 195 (2020).

95. Rhyou, H.-I. & Nam, Y.-H. Association between cognitive function and asthma in adults. Annals of Allergy, Asthma & Immunology 126, 69–74 (2021).

96. Ray, M., Sano, M., Wisnivesky, J.P., Wolf, M.S. & Federman, A.D. Asthma control and cognitive function in a cohort of elderly adults. Journal of the American Geriatrics Society 63, 684–691 (2015).

97. Alvarez, J.I., Cayrol, R. & Prat, A. Disruption of central nervous system barriers in multiple sclerosis. Biochimica et Biophysica Acta (BBA)-Molecular Basis of Disease 1812, 252–264 (2011).

98. Krupp, L.B., et al. International Pediatric Multiple Sclerosis Study Group criteria for pediatric multiple sclerosis and immune-mediated central nervous system demyelinating disorders: revisions to the 2007 definitions. Multiple Sclerosis Journal 19, 1261–1267 (2013).

99. Huda, S., et al. Neuromyelitis optica spectrum disorders. Clinical Medicine 19, 169 (2019).

100. Kim, W., Kim, S.-H., Huh, S.-Y. & Kim, H.J. Brain abnormalities in neuromyelitis optica spectrum disorder. Multiple sclerosis international 2012(2012).

101. Lancaster, E. The diagnosis and treatment of autoimmune encephalitis. Journal of Clinical Neurology 12, 1–13 (2016).

102. Wartolowska, K., et al. Structural changes of the brain in rheumatoid arthritis. Arthritis & Rheumatism 64, 371–379 (2012).

103. Kozora, E. & Filley, C.M. Cognitive dysfunction and white matter abnormalities in systemic lupus erythematosus. Journal of the International Neuropsychological Society 17, 385–392 (2011).

104. Appenzeller, S., et al. Longitudinal analysis of gray and white matter loss in patients with systemic lupus erythematosus. Neuroimage 34, 694–701 (2007).

105. Rosenberg, G.A. Inflammation and white matter damage in vascular cognitive impairment. Stroke 40, S20–S23 (2009).

106. Raj, D., et al. Increased white matter inflammation in aging-and Alzheimer’s disease brain. Frontiers in molecular neuroscience 10, 206 (2017).

107. Gerdts, E., et al. Correlates of left atrial size in hypertensive patients with left ventricular hypertrophy: the Losartan Intervention For Endpoint Reduction in Hypertension (LIFE) Study. Hypertension 39, 739–743 (2002).

108. Eshoo, S., Ross, D.L. & Thomas, L. Impact of mild hypertension on left atrial size and function. Circulation: Cardiovascular Imaging 2, 93–99 (2009).

109. Sanfilippo, A.J., et al. Atrial enlargement as a consequence of atrial fibrillation. A prospective echocardiographic study. Circulation 82, 792–797 (1990).

110. Saheera, S. & Krishnamurthy, P. Cardiovascular Changes Associated with Hypertensive Heart Disease and Aging. Cell Transplant 29, 963689720920830 (2020).

111. Hiraiwa, H., et al. Clinical significance of spleen size in patients with heart failure. European Heart Journal 42, ehab724. 0756 (2021).

112. Ormazabal, V., et al. Association between insulin resistance and the development of cardiovascular disease. Cardiovascular diabetology 17, 1–14 (2018).

113. Shah, A., Mehta, N. & Reilly, M.P. Adipose inflammation, insulin resistance, and cardiovascular disease. Journal of Parenteral and Enteral Nutrition 32, 638–644 (2008).

114. Boudina, S. & Abel, E.D. Diabetic cardiomyopathy, causes and effects. Reviews in Endocrine and Metabolic Disorders 11, 31–39 (2010).

115. Horton, W.B. & Barrett, E.J. Microvascular dysfunction in diabetes mellitus and cardiometabolic disease. Endocrine reviews 42, 29–55 (2021).

116. Kibel, A., et al. Coronary microvascular dysfunction in diabetes mellitus. Journal of International Medical Research 45, 1901–1929 (2017).

117. Fuentes-Antrás, J., et al. Targeting metabolic disturbance in the diabetic heart. Cardiovascular Diabetology 14, 1–11 (2015).

118. Wagner, R., et al. Metabolic implications of pancreatic fat accumulation. Nat Rev Endocrinol 18, 43–54 (2022).

119. Yaney, G.C. & Corkey, B.E. Fatty acid metabolism and insulin secretion in pancreatic beta cells. Diabetologia 46, 1297–1312 (2003).

120. Dludla, P.V., et al. Pancreatic beta-cell dysfunction in type 2 diabetes: Implications of inflammation and oxidative stress. World J Diabetes 14, 130–146 (2023).

121. Kocaturk, E., Kar, E., Kusku Kiraz, Z. & Alatas, O. Insulin resistance and pancreatic beta cell dysfunction are associated with thyroid hormone functions: A cross-sectional hospital-based study in Turkey. Diabetes Metab Syndr 14, 2147–2151 (2020).

122. Meeks, K.A.C., Adeyemo, A. & Agyemang, C. Beta-cell dysfunction and insulin resistance in relation to abnormal glucose tolerance in African populations: can we afford to ignore the diversity within African populations? BMJ Open Diabetes Res Care 10(2022).

123. Bonora, E., et al. Insulin resistance and beta-cell dysfunction in newly diagnosed type 2 diabetes: Expression, aggregation and predominance. Verona Newly Diagnosed Type 2 Diabetes Study 10. Diabetes Metab Res Rev 38, e3558 (2022).

124. Whalen, R., Carter, D. & Steele, C. Influence of physical activity on the regulation of bone density. Journal of biomechanics 21, 825–837 (1988).

125. Zhao, Q., Wang, J., Spiller, W., Bowden, J. & Small, D.S. Two-sample instrumental variable analyses using heterogeneous samples. Statistical Science 34, 317–333 (2019).

126. Cui, R., et al. Improving fine-mapping by modeling infinitesimal effects. bioRxiv, 2022.2010. 2021.513123 (2022).

127. Xue, H., Shen, X. & Pan, W. Causal Inference in Transcriptome-Wide Association Studies with Invalid Instruments and GWAS Summary Data. Journal of the American Statistical Association, 1–27 (2023).

128. Tseng, W.Y., Su, M.Y. & Tseng, Y.H. Introduction to Cardiovascular Magnetic Resonance: Technical Principles and Clinical Applications. Acta Cardiol Sin 32, 129–144 (2016).

129. Pennell, D.J. Cardiovascular magnetic resonance. Circulation 121, 692–705 (2010).

130. Bai, W., et al. Automated cardiovascular magnetic resonance image analysis with fully convolutional networks. Journal of Cardiovascular Magnetic Resonance 20, 1–12 (2018).

131. Bai, W., et al. Recurrent neural networks for aortic image sequence segmentation with sparse annotations. International conference on medical image computing and computer-assisted intervention, 586–594 (2018).

132. Miller, K.L., et al. Multimodal population brain imaging in the UK Biobank prospective epidemiological study. Nat Neurosci 19, 1523–1536 (2016).

133. Zhao, B., et al. Heritability of regional brain volumes in large-scale neuroimaging and genetic studies. Cerebral Cortex 29, 2904–2914 (2019).

134. Zhao, B., et al. Large-scale GWAS reveals genetic architecture of brain white matter microstructure and genetic overlap with cognitive and mental health traits (n= 17,706). Molecular psychiatry 26, 3943–3955 (2021).

135. Avants, B.B., et al. A reproducible evaluation of ANTs similarity metric performance in brain image registration. Neuroimage 54, 2033–2044 (2011).

136. Jahanshad, N., et al. Multi-site genetic analysis of diffusion images and voxelwise heritability analysis: A pilot project of the ENIGMA–DTI working group. Neuroimage 81, 455–469 (2013).

137. Kochunov, P., et al. Multi-site study of additive genetic effects on fractional anisotropy of cerebral white matter: comparing meta and megaanalytical approaches for data pooling. Neuroimage 95, 136–150 (2014).

138. Glasser, M.F., et al. A multi-modal parcellation of human cerebral cortex. Nature 536, 171–178 (2016).

139. Ji, J.L., et al. Mapping the human brain’s cortical-subcortical functional network organization. Neuroimage 185, 35–57 (2019).

140. Bowden, J., et al. Improving the accuracy of two-sample summary-data Mendelian randomization: moving beyond the NOME assumption. International journal of epidemiology 48, 728–742 (2019).

