## Supplementary material for "Inferring Multi-Organ Genetic Causal Connections using Imaging and Clinical Data through Mendelian Randomization": supp_information

**This PDF file includes:**

Supplementary Note
Legends for Tables S1 to S6

**Other Supplementary Materials for this manuscript include the following:**

Tables S1 to S6 (.xlsx) (available in a zip file)

**Supplementary Note**

Heritability of multi-organ imaging biomarkers

It has been shown that the CMR and brain MRI exists certain heritability by many twin and family studies[1-3]. However, the level of heritability might differ with respect to different heart or brain regions. For example, large brain volumes and cortical regions involved in language, executive function, and emotional regulations are usually consistently high in heritability[1,4]. Most of the brain structural MRI traits has relatively strong heritability, ranging from 0.6 to 0.8[4] compared with 0.2 to 0.6 heritability of brain functional connectivity[5]. Cardiac MRI traits are also heritable[6], such as ascending aortic[7] and left ventricular strcuture[8]. The heritability of all abdominal IDPs is significant, which suggests that genetic differences account for a substantial portion of the variation among individuals. The inclusion of height and BMI as additional covariates has minimal impact on heritability. More detailed information about abdominal IDPs can be found in Liu, et al. [9]. All skeletal proportions are highly heritable (0.4-0.5) and the range in twin studies is between 0.4 and 0.8[10].

### References


1. Schmitt, J.E., *et al.* Review of twin and family studies on neuroanatomic phenotypes and typical neurodevelopment. *Twin Res Hum Genet* **10**, 683-694 (2007).
2. Batouli, S.A., Trollor, J.N., Wen, W. & Sachdev, P.S. The heritability of volumes of brain structures and its relationship to age: a review of twin and family studies. *Ageing Res Rev* **13**, 1-9 (2014).
3. Lopes, M.C., Andrew, T., Carbonaro, F., Spector, T.D. & Hammond, C.J. Estimating heritability and shared environmental effects for refractive error in twin and family studies. *Invest Ophthalmol Vis Sci* **50**, 126-131 (2009).
4. Jansen, A.G., Mous, S.E., White, T., Posthuma, D. & Polderman, T.J. What twin studies tell us about the heritability of brain development, morphology, and function: a review. *Neuropsychol Rev* **25**, 27-46 (2015).
5. Foo, H., *et al.* Genetic influence on ageing-related changes in resting-state brain functional networks in healthy adults: A systematic review. *Neurosci Biobehav Rev* **113**, 98-110 (2020).
6. Pirruccello, J.P., *et al.* Analysis of cardiac magnetic resonance imaging in 36,000 individuals yields genetic insights into dilated cardiomyopathy. *Nat Commun* **11**, 2254 (2020).
7. Tcheandjieu, C., *et al.* High heritability of ascending aortic diameter and trans-ancestry prediction of thoracic aortic disease. *Nat Genet* **54**, 772-782 (2022).
8. Jin, Y., *et al.* Heritability of left ventricular structure and function in Caucasian families. *Eur J Echocardiogr* **12**, 326-332 (2011).
9. Liu, Y., *et al.* Genetic architecture of 11 organ traits derived from abdominal MRI using deep learning. *Elife* **10**, e65554 (2021).
10. Chatterjee, S., Das, N. & Chatterjee, P. The estimation of the heritability of anthropometric measurements. *Applied Human Science* **18**, 1-7 (1999).

**Legends for Tables S1 to S6** (All tables can be found in a zip file).

**Table S1. ID of Finngen clinical endpoints used in the study.**

**Table S2. ID of imaging biomarkers used in the study.**

**Table S3. Estimates of causal genetic links between brain imaging biomarkers and clinical endpoints.**

**Table S4. Estimates of causal genetic links between heart imaging biomarkers and clinical endpoints.**

**Table S5. Estimates of causal genetic links between abdominal imaging biomarkers and clinical endpoints.**

**Table S6. Estimates of causal genetic links between skeleton imaging biomarkers and clinical endpoints.**
